# Mortality outcomes after removal from end-of-life registers: A prospective cohort-controlled study

**DOI:** 10.64898/2026.03.15.26348422

**Authors:** Amy Thompson, Emily Heyting, Vijay Klaire, Jonathan Lampitt, Baldev Singh, Wolverhampton Digital Health Primary Care Research Network, Emma Parry

## Abstract

**Background:** Earlier identification and registration of people in the last year of life improves care quality and outcomes in general practice. However, there is little evidence on patients who subsequently no longer require end-of-life registration, nor on the safety or outcomes of de-registration following clinical review.

**Aim:** To determine the prevalence, safety, and prognostic validity of GP-led removal from the end-of-life register (EOL_R) using a systematic digital review process.

**Design and Setting:** Observational cohort study in eight practices in Wolverhampton, UK, using a whole-population integrated primary and secondary care dataset.

**Method:** All adults on the EOL_R were systematically reviewed using a digital end-of-life pathway (PRADA) incorporating robotic process analysis of recognised end-of-life care markers. GPs recorded a binary decision to retain or remove patients from the register. Mortality outcomes were compared with those retained on the EOL_R, a tightly propensity-matched cohort not on the register, and the residual general population over 15 months’.

**Results:** Of 422 registered patients, 33 (7.8%) were removed following GP assessment. One-year survival in the removed group was statistically indistinguishable from the propensity-matched control cohort, and survival was significantly higher versus those retained on the EOL_R (60.4%, p<0.001). Removal demonstrated a negative predictive value for mortality of 90.9%.

**Conclusion:** GP-led removal from the end-of-life register can be undertaken safely and identifies a distinct group with substantially better prognosis. Digital systems that support systematic review, documentation, and follow-up should be incorporated into routine practice and reflected in national guidance and the Quality and Outcomes Framework.

**Statement boxes:** *What is known:* Earlier identification and registration of people in the last year of life improves care coordination and outcomes. However, little is known about patients who subsequently no longer require end-of-life registration, or whether removal following clinical review is associated with adverse outcomes.

*What this study adds:* In a whole-population primary care cohort, General Practitioners removed 33 patients (7.8% of those registered) from the end-of-life register following structured clinical review. One-year survival in this group was equivalent to a tightly matched cohort not on the register, and survival was substantially higher than among patients retained on the register. A simple robotic based review process can prompt systematic reassessment, capture GP clinical judgement, and enable prospective monitoring following removal.

*Implications:* End-of-life registration should be treated as a dynamic process requiring ongoing clinical review. Digital systems can support safe removal from registers by documenting decisions and embedding follow-up for patients whose prognosis remains uncertain. Evidence-based guidance and governance processes for a review process are needed to ensure people are not retained on registers unnecessarily.

*How this fits in:* End-of-life registers are intended to support proactive care for people in their last year of life, yet there is little evidence about patients who later stabilise and may no longer require registration. In eight UK practices, a systematic GP review supported by a digital end-of-life pathway identified a small but clinically important group suitable for removal, without adverse mortality outcomes. Beyond improving register accuracy, structured review creates opportunities for meaningful dialogue with patients, shared reassessment of care goals, and appropriate de-escalation of end-of-life labelling. Embedding routine, structured review with documented decisions and follow-up may also reduce unnecessary clinical workload and strengthen governance in primary care.

*Novelty Statement:* Systematic GP review of end-of-life registers, as stipulated in GMC guidance, is rarely evaluated. We propose a digitally driven, systematic and dynamic clinical-governance-led approach to register review.

## INTRODUCTION

Promoting earlier identification and end-of-life registration (EOL_R) for people who may be in their last year of life improves anticipatory care, supports better coordination across services, and is fundamental to improving care experiences as described in the NHS 10-year plan (1). This is aligned with national policy and guidance, and sits within wider ambitions for NHS digitally enabled care delivery (2,3). Digital systems could better utilise rich data, integrated between providers, to capture whole populations for effective risk stratification. This data can be deployed in real-time for clinical decision making, thus identifying those in need of end-of-life (EOL) processes and driving cohesive care between fragmented providers. Despite this, current digital EOL systems often have limited scalability, rely on complex modelling that is difficult to operationalise in routine practice, or report insufficient outcome evaluation (3,4).

Within our local health economy, as part of a broader digital health ambition, we developed a specific EOL digital care pathway, the Proactive Risk-Based and Data-Driven Assessment of Patients at the End of Life (PRADA), combining robotic process and machine learning techniques with clinical assessment (3, 5-10) to support earlier identification and registration for EOL care (5-10). Most recently, we demonstrated the efficacy of a robotic process-driven EOL “BOT” to identify patients for clinical validation by their GP for inclusion on the EOL register (8). Clinicians are generally more confident and accurate in identifying people who are clearly in the last phases of life or those not approaching end of life, with greater uncertainty in intermediate prognostic trajectories (9-12), where patient-centred discussion and shared decision-making remain essential and cannot be reduced to risk scores alone. Accordingly, we have developed a “digital EOL_R tracker” to ensure that these “grey” cases are regularly reviewed (8, 13).

With mounting focus on proactive case-finding for end-of-life registration (EOL_R), a related but overlooked area is the review of patients already on the register. Professional guidance emphasises ongoing review and reassessment of care needs towards the end of life, including changes over time (14). Inevitably this will result in a cohort of patients who stabilise and no longer require registration, but little is known about the impact of de-registration for patients, the frequency of removal from the register, or outcomes following removal.

The study therefore used PRADA to support a systematic GP review of all patients on an EOL_R, to determine the prevalence of removal following GP clinical judgement and to compare subsequent mortality outcomes across defined cohorts.

## METHODS

### Setting

The study was conducted across eight general practices in Wolverhampton, UK, a multiethnic city with high levels of socioeconomic deprivation.

### Research question

What is the subsequent mortality of patients removed from the end-of-life register following GP clinical review?

### Main Outcome

All-cause mortality over the study period.

### Study design

This was a prospective, cohort-controlled study. From the total registered population aged 18 years and over (n=39,079), all patients on the end-of-life register (EOL_R) were included, with no additional inclusion or exclusion criteria. A tightly matched control cohort was constructed by propensity matching all patients on the EOL_R at baseline (n=422) 1:1 with patients not on the register and without any recorded end-of-life care processes, matched exactly for age, sex, ethnicity, index of multiple deprivation, and multimorbidity (≥3 long-term conditions). Each patient on the EOL_R at baseline was reviewed by their GP, who recorded a binary “yes” or “no” decision to either retain or remove the patient from the register. Any subsequent clinical actions arising from the GP review, including discussion and agreement with the patient, were undertaken as part of usual clinical care. This resulted in four cohorts: patients retained on EOL_R; patients removed from EOL_R; a propensity-matched control cohort; and the residual registered population. All cohorts were followed prospectively for mortality. The study duration was 15 months, ensuring a minimum follow-up of 6 months after the final GP review.

### The PRADA system

Information on all patients was presented to GPs in a dedicated digital system built on an integrated health data resource. As well as general information on unscheduled care and care complexity, this displayed any end-of-life process recorded across the wider health economy. The five process markers of EOL care were the Surprise Question outcome (“No, I would not be surprised if the person died in the next year”), Electronic Palliative Care Coordinating System (EPaCCS) documentation, Recommended Summary Plan for Emergency Treatment and Care (ReSPECT) documentation, any advanced EOL care plans and any coded contact with the hospital Specialist Palliative Care team. These processes were combined in a simple robotic process assessment termed “BOT”. The presence or absence of any single EOL process was termed “BOT positive” or “BOT negative”, respectively, with the total process number also displayed (8). The GP’s clinical decision was recorded with a date and time stamp. Any patients identified for removal from the EOL register were flagged and their continued status monitored to verify removal had occurred. As a “fail safe” process, all removed patients were retained in a surveillance list “the digital EOL_R tracker” (8) with a digitally mandated review after 6 months, or earlier, as set by the GP. In this study, the BOT was used solely to support systematic review of patients already registered on the EOL_R, and not to identify new patients for inclusion on the register.

### Data

The Wolverhampton Integrated Clinical Data Set links demographic, clinical and emergency activity data from primary care, hospital, and community services data under General Data Protection Regulation. The 5 process markers of an end-of-life pathway described above were ascertained from that combined resource. Frailty was assessed by the Rockwood score, or in its absence the Electronic Frailty Index, and then risk categorised into a binary variable of ‘moderate / severe frailty’ versus “non-frail”. Mortality was determined from hospital mortality statistics and rolling NHS Strategic Tracing Service checks.

### Statistical method

All data were analysed on IBM SPSS version 29. When comparing independent groups, multiple group means were tested by one way analysis of variance, and the Chi-square test was used for the difference between proportions. Post-hoc 2 group statistical comparisons were only undertaken if that was significant. Survival analysis was by Cox’s regression. Results are presented as the mean ± standard deviation (SD) or as numbers with percentages. Statistical significance of all tests applied was taken at p<0.05.

### Research check list

Strengthening the Reporting of Observational Studies in Epidemiology for cohort studies (15).

### Ethical considerations

The study did not involve selection criteria, randomisation, nor any intervention that ought not to have otherwise happened. Ethics approval and informed consent was not deemed necessary for this study according to our Institutional Review Board (Royal Wolverhampton NHS Trust Research and Development Department). The study was considered a clinical governance driven quality improvement project using innovative digital healthcare methodologies.

### Patient consent

Not applicable

### Patient and Public Involvement

For this proof-of-concept phase, preliminary dialogue on the intended use of integrated clinical data in digital health care methodologies occurred with the patient advisory and liaison group of Royal Wolverhampton NHS Trust.

## RESULTS

Across eight practices, the registered adult population was 39,079. Among 422 patients on the EOL_R, GPs identified 33 (7.8%) for removal following clinical review.

**Table 1** summarises the cohort characteristics. In post-hoc analysis, compared with the tightly propensity-matched control cohort (n=354), patients removed from the EOL_R had a higher prevalence of nursing home residence, but did not differ on other matches variables, including mortality (9.1% vs 7.1%, not significant). Compared with patients retained on EOL_R, those removed were younger, had fewer urgent care events, were less likely to reside in a nursing home, and had significantly lower mortality (7.8% vs 39.6%, p<0.001).

**Table 1.** Demographic, clinical and end-of-life parameters in the 4 cohorts derived from the whole population studied (n=39,079). Statistical comparison was by one way analysis of variance or the Chi square test for groups means or the difference between proportions respectively. Post-hoc 2 group comparisons were as indicated, with single or triple symbols denoting p<0.05 or p<0.001. EOL = end-of-life, EPL_R = end-of-life register, A/E = Accident and Emergency department, NEA = non-elective admissions. Index of Multiple Deprivation is a UK area-level measure of socioeconomic deprivation, derived from residential postcode, with higher scores indicating greater deprivation.

|  | Unselected residual general population | Propensity matched control cohort for those originally on the EOL_R | EOL_R de-escalated following GP assessment. Post-hoc comparison to group 2 | EOL_R retained following GP assessment. Post-hoc comparison to group 3. | 4 group statistical comparison |
| --- | --- | --- | --- | --- | --- |
| Cohort number | 1 | 2 | 3 | 4 |  |
| Originally end-of-life registered (n = 422) | No | No | Yes | Yes |  |
| Number (of 39,079 total) | 38,303 (98.0%) | 354 (0.9%) | 33 (0.1%) | 389 (1.0%) |  |
| Any end-of-life process | 546 (1.4%) | 0 | 0 | 344 (88.4%) ^^^ | X2 = 13,107.7, $p < 0.001$ |
| Died by end of study period | 437 (1.1%) | 25 (7.1%) | 3 (9.1%) | 154 (39.6%) ^^^ | X2 = 3,732, $p < 0.001$ |
| Days to Death in those that died | 251 +/- 128 (0 - 471) | 234 +/- 132 (0 - 457) | 206 +/- 123 (110 - 345) | 211 +/- 128 (23 - 468) | F = 3.8, $p < 0.01$ |
| Mortality in those with no EOL process | 293 / 37,757 (0.8%) | As above | As above | 16 / 45 (35.6%) ^^^ | X2 = 803.4, $p < 0.001$ |
| Mortality in those with any EOL process | 144 / 546 (26.4%) | - | - | 138 / 344 (40.1%) | X2 = 18.4, $p < 0.001$ (2 groups) |
| Age (years) | 47.2 +/- 18.1 (18-104) | 77.2 +/- 11.9 (24-98) | 75.2 +/- 14.8 (42-96) | 80.6 +/- 13.0 (23 - 103) ^ | F = 778.3, $p < 0.001$ |
| Gender (Male) | 19,578 (51.1%) | 157 (44.4%) | 18 (54.5%) | 163 (41.9%) | X2 = 19.5, $p < 0.001$ |
| Ethnicity (White) | 21,703 (55.4%) | 300 (84.7%) | 25 (75.8%) | 333 (85.6%) | X2 = 268.7, $p < 0.001$ |
| Index of multiple deprivation | 28.6 +/- 16.3 (2.1 - 71.8) | 19.9 +/- 11.1 (4.1 - 71.8) | 18.4 +/- 13.6 (7.2 - 50.1) | 19.2 +/- 10.7 (6.6 - 56.7) | F = 80.2, $p < 0.001$ |
| >= 3 A/E attendances (12 months) | 456 (1.2%) | 8 (2.3%) | 1 (3.0%) | 29 (7.5%) | X2 = 124.8, $p < 0.001$ |
| >= 3 NEA (12 months) | 243 (0.6%) | 6 (1.7%) | 1 (3.0%) | 41 (10.5%) | X2 = 517.9, $p < 0.001$ |
| Either >= 3 A/E or >= 3 NEA (12 months) | 563 (1.5%) | 11 (3.1%) | 1 (3.0%) | 59 (15.2%) ^ | X2 = 458.0, $p < 0.001$ |
| >= 3 Co-morbidities | 3,948 (10.3%) | 252 (71.2%) | 26 (78.8%) | 290 (74.6%) | X2 = 2,946.3, $p < 0.001$ |
| Frailty (moderate or severe) | 4,068 (10.6%) | 226 (63.8%) | 24 (72.7%) | 325 (83.5%) | X2 = 2,996.1, $p < 0.001$ |
| Nursing home resident | 262 (0.7%) | 5 (1.4%) | 5 (15.2%) *** | 226 (58.1%) ^^^ | X2 = 10,140.3, $p < 0.001$ |

Notably, all patients removed from the register had no recorded end-of-life process markers and were therefore BOT-negative, indicating that they had been registered on the EOL_R prior to the BOT process.. In the original EOL_R cohort (n=422), 78/422 (18.4%) were BOT-negative; 33/78 (42.3%) were removed, while 45 (57.7%) were retained. Mortality over the 15-month follow-up period among BOT-negative patients who were retained was higher than among those removed (16 / 45 (35.6%) vs 3/33 (9.1%), X2=7.24, p=0.007) **(Table 1)**, and was not statistically distinguishable from mortality among those retained on EOL_R with any recorded EOL process marker (138/344 (40.1%), X2 = 0.35, not significant).

Furthermore, in the residual population not on the EOL_R **(Table 1)**, 546/38,303 (1.4%) had a recorded EOL process marker and had significantly higher mortality (144/546, 26.4%) than the removed cohort (X2 = 4.9, p<0.05). GP-assessed retention on the EOL_R captured 154/157 deaths among those originally registered (sensitivity 98.1%), whilst removal had a negative predictive value for mortality of 90.9% (30/33 alive). **Figure 1** shows cumulative survival across cohorts.

**Figure 1.**
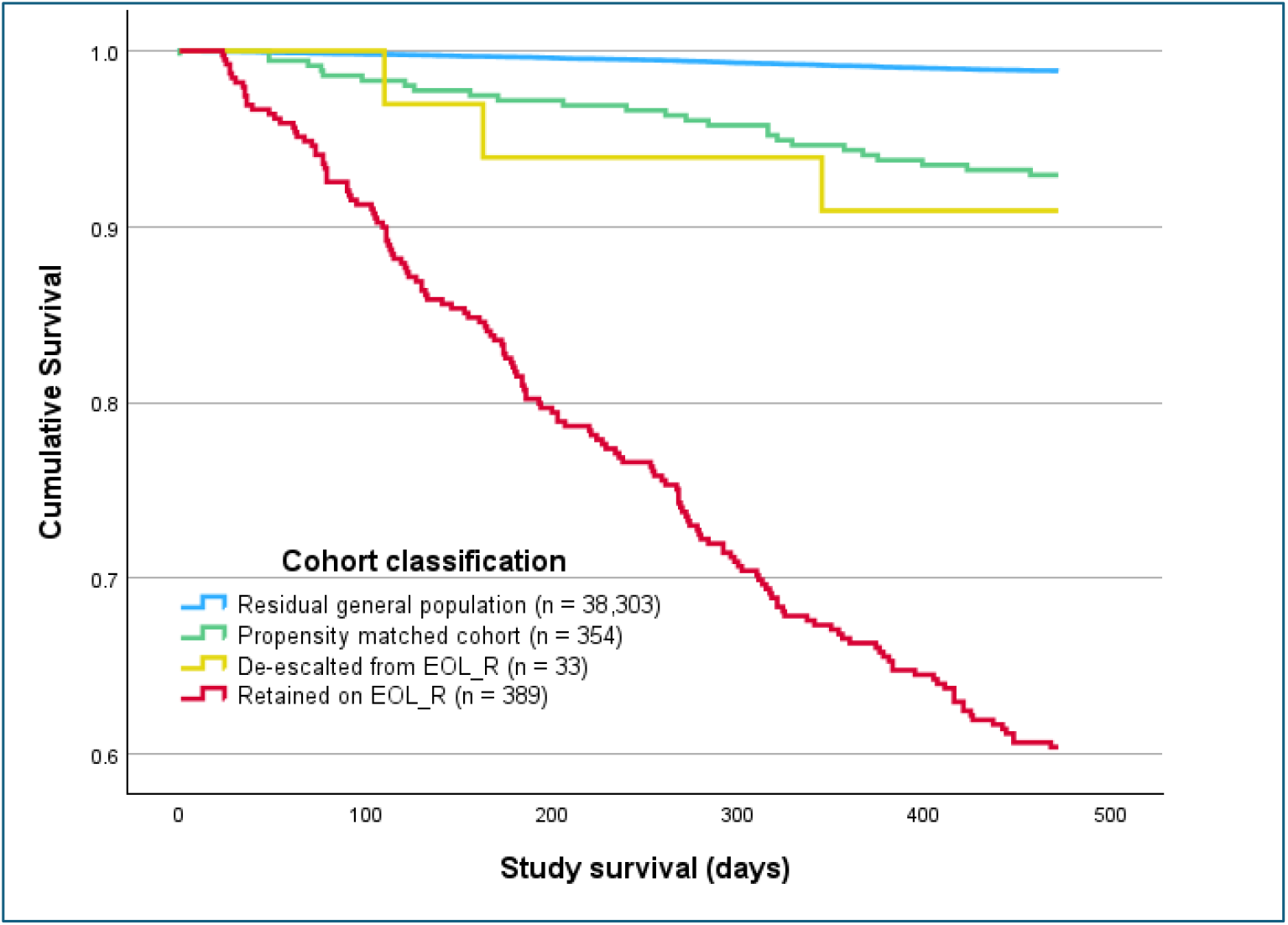
Survival analysis by Cox’s regression over the study period amongst the 4 cohorts derived from the whole population studied (n=39,079) as defined in the Table.

## DISCUSSION

### Summary

In this whole-population cohort study, systematic GP review of all patients on an end-of-life register identified a small but important subgroup (7.8%) for removal following clinical review. Subsequent mortality among those removed was statistically indistinguishable from a tightly matched control cohort, and significantly lower (p<0.001) than among patients retained on the register, even when sub-selecting those retained on the EOL_R with no EOL process markers (p<0.001), and lower than those in the general population with a recorded EOL process marker (p<0.001). These findings support end-of-life registration as a dynamic process requiring routine reassessment, with digital systems able to prompt structured review, record decisions, and enable follow-up.

### Strengths and limitations

Strengths include the use of an integrated multi-provider dataset applied to an unselected primary care population; prospective ascertainment of mortality; a tightly propensity-matched control cohort; and explicit capture of GP clinical judgment at the point of decision-making. The PRADA system also incorporated a practical governance mechanism, retaining removed patients on a surveillance list with mandated review.

Limitations include small absolute numbers in the removed cohort and few deaths, which reduces precision around mortality estimates in this subgroup. The study evaluates mortality as the primary outcome; it does not directly measure other potential consequences of removal (for example, access to services or patient-reported outcomes). This study did not capture data on the specific clinical reasons underpinning decisions to remove patients from the EOL_R. While GP judgement was recorded prospectively and outcomes were followed longitudinally, the study did not categorise the clinical rationale for de-escalation. Future work should explore the reasons for removal, such as stabilisation, diagnostic revision, or changes in patient preference, to further inform guidance and decision support.

### Comparison with existing literature

It is recommended that patients on an EOL_R are reviewed as circumstances change for continued inclusion or removal as circumstances change (14). Such review is also reflected in UK primary care incentive frameworks, although less visible in wider national EOL guidance (2). Inevitably, some patients on an EOL_R will stabilise or improve, meaning that removal from the register may be clinically appropriate, even though overall prognosis may remain guarded. We are unaware of any prior publications describing the frequency of removal or outcomes following removal.

Clinical judgment is often treated as subjective and not always explicitly incorporated into digital models, despite the fact that EOL prognostication and EOL_R inclusion commonly rely on clinical assessment in practice (9-12). A recognised framework exists for integrating clinical judgement with probabilistic reasoning (17). Alongside this, there is sustained emphasis on improving identification for EOL registration because many people who die are not registered; where early identification is combined with effective anticipatory planning, benefits include improved coordination and reduced crisis-driven care (2). Digital prediction methodologies can support case-finding, and evidence suggests their accuracy can be strengthened when combined with clinician judgment (5-10). However, prognostication remains inherently uncertain, particularly for intermediate trajectories, and decisions reflect multiple factors including timeliness, patient wishes and their broader needs (7, 16). For these reasons, removal from an EOL_R could reasonably be considered a high-stakes decision, with potential unintended outcomes if undertaken without governance (for example, missed opportunities for anticipatory planning or avoidable unscheduled care).

In this study, 7.8% of patients originally on the EOL_R were removed following GP clinical judgement; this proportion is likely to vary by setting and case mix. More importantly, removal identified a group with substantially better subsequent survival than those retained on the register, and similar survival to a tightly matched control cohort not on the register. Although mortality among removed patients was lower than among those retained, it was not negligible (9.1%), reflecting the underlying age and morbidity of this group. To support clinical governance and maintain a safety net, the PRADA system retains removed patients on a digital surveillance list, known as the “digital end-of-life register tracker”, with mandated review at 6 months or sooner if clinically indicated (8). Taken together, these findings suggest that GP review adds meaningful prognosis discrimination within the EOL_R population when making decisions to retain or remove.

PRADA is a growing programme of work describing digitally enabled end-of-life care delivery at whole-population scale using routinely available clinical and activity data, with clinician interaction embedded within workflow, and utilising a combination of robotic process and machine learning digital methodologies (3,5-10), with a specific focus on the quality of the primary care EOL_R (both for case identification and now also defining systematic processes for EOL_R review). While PRADA uses both robotic process and machine learning methods for risk stratification and mortality prediction, this study focuses on the robotic-process EOL_R “BOT” based on the presence of key EOL process markers. In this study, 78/422 (18.4%) of those originally on the EOL_R were BOT-negative; this group included all patients removed, alongside a smaller subgroup who were retained and had substantially higher mortality. A further subgroup in the general population was BOT-positive and had comparatively high mortality, supporting the BOT as a pragmatic signal for review. We are not aware of any other EOL digital pathways that explicitly combine (i) systematic review of the existing EOL_R, (ii) structured capture of clinical judgement, and (iii) prospective outcome surveillance following removal into an “end-to-end” approach.

### Implications

In the light of these findings and existing professional guidance, a structured, well-governed, and digitally led approach to reviewing the end-of-life register - including the option of removal where appropriate - should be normalised within routine practice (2,3,14). Although national guidance emphasises the importance of ongoing review as circumstances change, it provides little direction on the optimal timing or structure of reassessment. Digitally supported systems that prompt review, document clinical decisions, and embed flexible follow-up may help operationalise this expectation in everyday primary care, improving both safety and confidence in dynamic register management (3,8).

Organisations focused on EOL care and the research community should test this approach in other populations, including evaluation of outcomes beyond mortality (for example, patient- and carer-centred outcomes). NICE has previously highlighted limitations in the evidence base for service models supporting identification and delivery of end-of-life care, and this area of register review and removal warrants explicit attention (18,19). At a time of intense pressure in UK general practice, commissioning and incentive frameworks should support not only earlier identification and inclusion on EOL registers, but also systematic review to maintain accuracy over time. System leaders and primary care record system suppliers should consider how these processes can be implemented at a national scale with a degree of immediacy, to mitigate fragmented and inefficient care, reduce avoidable hospital admissions and provide efficiency support and funding to primary care in their ambition to deliver better outcomes (1,3). It is our view that this digital approach at scale is readily achievable now.

### Summary

Using a digitally supported review process, GPs systematically reviewed all patients on their EOL_R and identified a cohort of patients requiring removal for clinical reasons. This cohort had a significantly lower subsequent mortality than patients retained on the register and survival similar to matched controls, supporting end-of-life registration as a dynamic process requiring structured review and governance.

## Data Availability

The corresponding / accountable author(s) will share anonymised study data upon reasonable request to, subject to approval of any proposal by the authors and our research governance body.

## Abbreviations used (List)

A/E: Accident and Emergency
BOT: Robotic Process Assessment of End-of-Life Care Processes
EHR / EPR: Electronic Patient Record
EOL: End of Life
EOL_R: End-of-Life Register
EPaCCS: Electronic Palliative Care Coordination System
GP: General Practitioner
IMD: Index of Multiple Deprivation
NEA: Non-Elective Admission
PRADA: Proactive Risk-Based and Data-Driven Assessment of Patients at the End of Life
ReSPECT: Recommended Summary Plan for Emergency Care and Treatment
SD: Standard Deviation

## Acknowledgements

The Wolverhampton Digital Heath Primary Care Research Network is a group of collaborating Wolverhampton GP practices implementing this programme in real time direct care, contributing to its development, clinical deployment, and continuous improvement, without which this ongoing innovation project would be impossible. The individual GPs are Drs I Adam, K Ahmed, J Burrell, AJ Cook, U Jaswal, A Koodaruth, C Madzima, G Malhi, G Pickavance, E Qureshi, M Sidhu. Contacts

